# A digital program to prevent falls and improve well-being in people living with dementia in the community: the KOKU-LITE feasibility Randomised Controlled Trial protocol

**DOI:** 10.1101/2024.07.17.24310446

**Authors:** Jaheeda Gangannagaripalli, Emma R.L.C Vardy, Emma Stanmore

## Abstract

**Introduction:** Around 885,000 people live with dementia in the UK of which around 50% experience a fall each year. ‘Keep On Keep Up’ (KOKU) is an NHS approved gamified, digital health program designed to maintain function and reduce falls through strength & balance exercises (FaME/OTAGO), and health literacy games. KOKU has been adapted to the needs of people living with Dementia (PLwD) in the community, known as KOKU-LITE. This trial aims to test the feasibility and acceptability of trial processes and usability of KOKU-LITE.

**Methods and analysis:** A two-arm, mixed methods, feasibility randomised controlled trial will be conducted. Participants aged ≥55 years meeting the eligibility criteria will be recruited from patient organisations across Greater Manchester, UK. Participants randomised into the intervention arm will receive 6 weeks KOKU-LITE program plus dementia specific falls prevention leaflet and participants randomised into the control arm will receive dementia specific falls prevention leaflet. Outcome measures include: recruitment rate, adherence, quality of life, participants’ Activities of Daily Living, physical activity levels, functional ability, lower limb strength, fear of falling, falls risk, mood, and user’s experience of the technology. Post-intervention interviews or focus groups with participants and health and social care professionals will explore feasibility of trial processes & technology and evaluate the usability and acceptability of the intervention respectively. Analyses will be descriptive.

**Ethics and dissemination:** This feasibility trial has been reviewed and received favourable ethical approval from Yorkshire & The Humber - Bradford Leeds Research Ethics Committee, Newcastle upon Tyne (REC reference 23/YH/0262). The findings of the study will be disseminated through peer-reviewed scientific journals, at conferences, publication on University of Manchester, Applied Research Collaboration Greater Manchester (ARC-GM) and KOKU websites.

**Trial registration number:** NCT06149702

**Strengths and Limitations of this study:**

1. This is one of the first studies to test the feasibility and acceptability of trial processes and usability of a gamified digital health program for falls prevention and to improve well-being in people living with dementia in the community known as KOKU-LITE.
2. Recruiting and retaining people living with dementia can be very challenging and therefore this feasibility study will explore and evaluate different strategies to recruit participants and estimate the time required for recruitment.
3. The barriers and facilitators identified in the recruitment and retention phase of the trial will help us to design a robust definitive trial.
4. We have involved people with lived experience of dementia extensively in the development of KOKU-LITE and will continue to do so in feasibility testing of KOKU-LITE to inform a larger study that will test effectiveness.
5. This is a feasibility trial and is not powered to determine the effectiveness of the intervention.

## BACKGROUND

It is estimated that over 885,000 people with dementia currently live in the UK with this figure anticipated to increase to 1.6 million by 2040 (1). The total cost of Dementia (including health and social care costs, and opportunity costs of unpaid care) in England in 2015 was £23 billion and is predicted to increase to over £80 billion by 2040 (2). Due to its increasing prevalence, Dementia is recognised as an international priority by the World Health Organisation (WHO) (3).

Falls are a public health concern and a common problem in older people (aged 65 years and over). One in three older adults fall each year (4,5) and those with cognitive impairment are more than twice as likely to fall compared to older adults without cognitive impairment (6–8). In addition, one in five falls causes serious injuries such as fractures or head injuries (4). The risk is compounded in older people living with Dementia (PLwD) due to additional risk factors (9). Falls are estimated to cost over £4 billion annually (10). Evidence related to interventions that are aimed at reducing falls for PLwD was found to be insufficient and inconclusive (11–13).

There is a growing interest in using digital technologies to support PLwD, and the use is recommended by WHO to promote peoples’ health and well-being (14). Utilising digital technology to enable older people living with dementia to live in their own home independently for longer, or to prevent falls has the potential to improve quality-of-life and provide cost savings to health and social care (15,16). However, there is a lack of robust evidence to suggest effective digital technologies for people with Dementia. Evidence also suggests that people living with Dementia are reluctant to use digital technologies due to the following barriers: Dementia severity, poor or lack of digital literacy, timing and disease progression, technology anxiety, system failures, digital divide, lack of access to or knowledge of how technology works, cognitive fatigue, and usability issues (17). Hence, it is crucial that digital interventions are designed in a user-centred manner for technologies to be acceptable (18–20). Digital technologies have the potential to assist relatives/carers of PLwD in predicting, detecting, monitoring, and prevention of falls in PLwD (21,22).

“Keep on Keep up (KOKU),” is a, tablet/iPad-based, digital gamified strength and balance exercise program specifically designed <u>with</u> older people <u>for</u> older people at risk of falls (23). KOKU supports older people to engage with simple, progressive falls prevention exercises; with the aim of helping people remain functionally active for longer and preventing falls. The strength and balance exercises are based on the evidence-based OTAGO and The Fitness and Mobility Exercise (FaME) Programme for community dwelling older adults (24,25) that have been shown to reduce falls by around a third. KOKU is General Data Protection Regulation (GDPR) compliant and has The Organisation for the Review of Care and Health Apps (ORCHA) (26) approval for National Health Service (NHS) use in the UK.

Following Medical Research Council complex intervention guidance (27), we have engaged with PLwD and carers of PLwD and co-developed KOKU to suit the needs of PLwD to make it dementia-friendly and more accessible by simplifying the language and other features (Supplementary appendix 1). This version is known as KOKU-LITE. This current project continues the involvement of PLwD and carers of PLwD in feasibility testing of KOKU-LITE to inform a larger study that will test effectiveness.

### Aim

To test the feasibility of the KOKU-LITE digital program with people living with Dementia (PLwD) aimed at preventing falls and improving well-being compared with control (a Dementia specific falls prevention leaflet).

### Objectives To

1) Explore study recruitment approaches for PLwD, their carers and Health and social care professionals (HSCPs) for the feasibility study.
2) Explore the training and support required for PLwD, their carers and HSCPs for the use of KOKU-LITE during the feasibility study.
3) Explore the usability and acceptability of the KOKU-LITE program.
4) Describe and determine the suitability of the outcome measures including providing data to permit estimation of effect size to be used in sample size calculations for a definitive trial.

## METHODS

### Study design

#### Intervention: KOKU-LITE program + a Dementia specific-falls prevention leaflet

##### Control: a Dementia specific-falls prevention leaflet

This feasibility randomised controlled trial is a 6-week study comparing the intervention to control in community dwelling PLwD. After assessing participants for eligibility, they will be allocated randomly to either intervention group or control group. Assessments will be performed at baseline and 6 weeks post follow up (figure 1).

**Figure 1.**
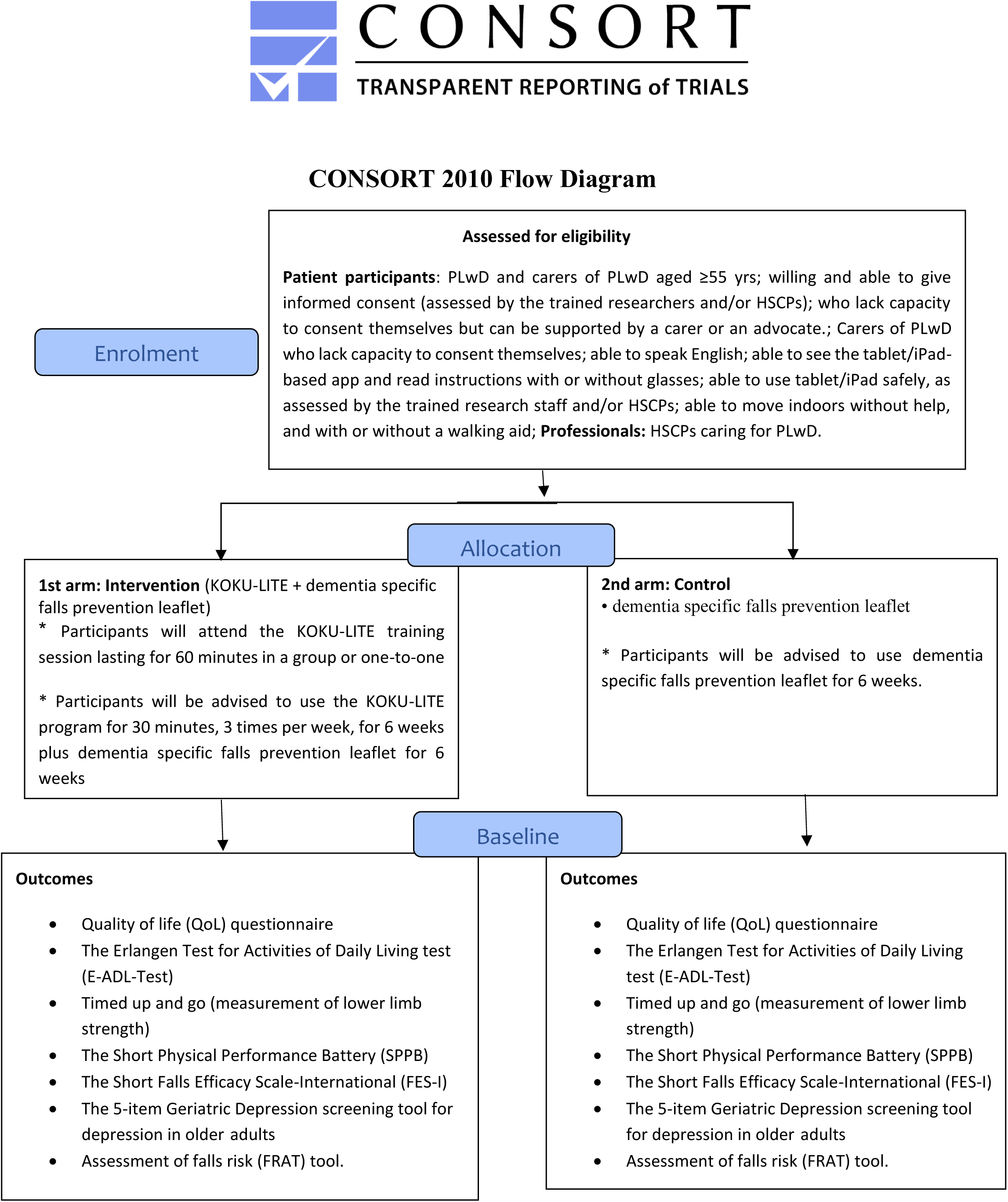
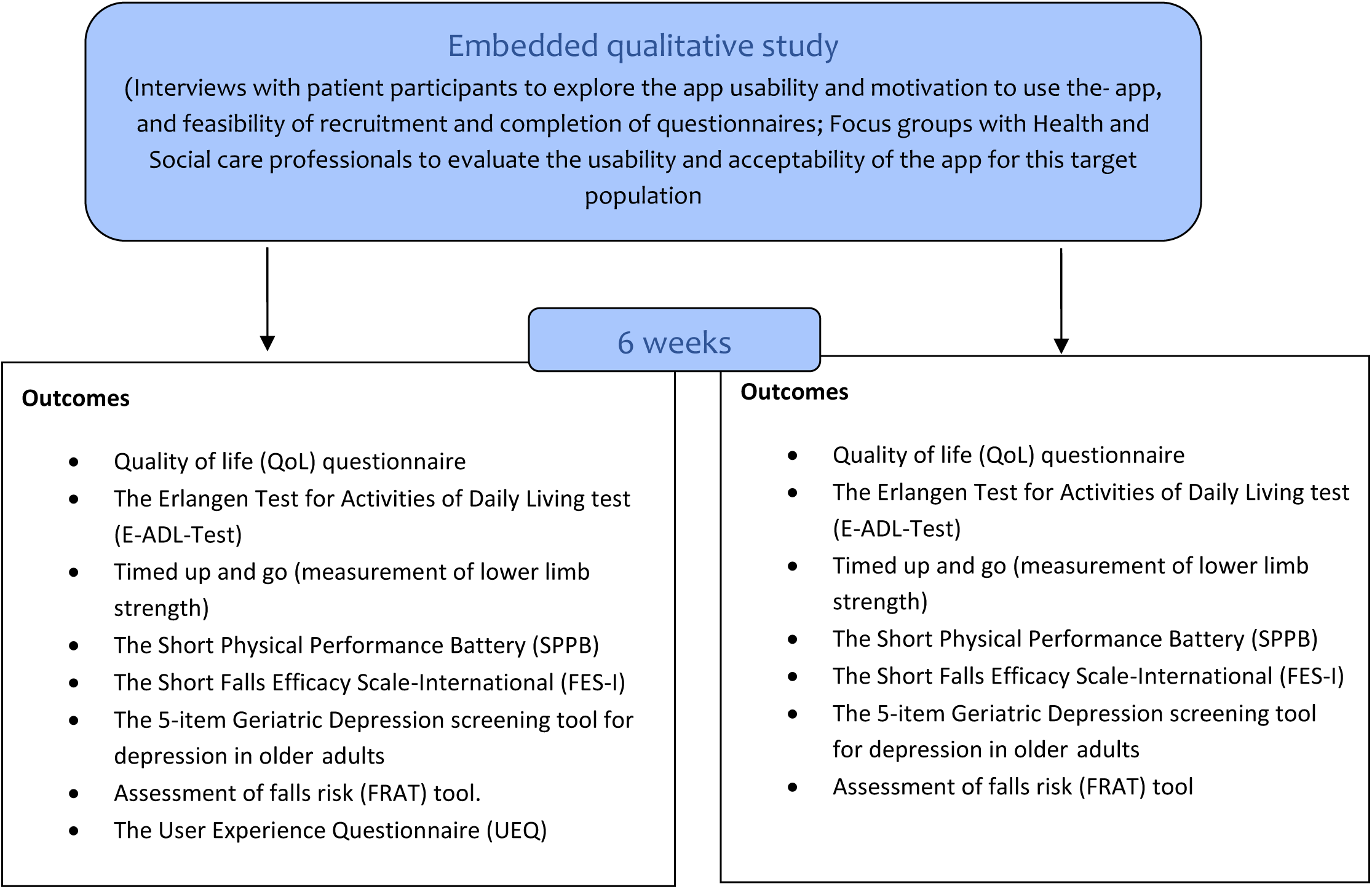

A purposive sample of participants who have consented to take part in the study will be invited to participate in post-intervention interviews. Exploratory data obtained from the interviews will increase understanding of the responses in the questionnaires about the usability and motivation to use KOKU-LITE, and feasibility of recruitment, data collection approaches and completion of outcome measures.

Patient recruitment has commenced in March 2024 and data collection will be completed by September 2024. The study will be reported in accordance with the Standard Protocol Items: Recommendations for Interventional Trials (SPIRIT) 2012 Statement for protocols of clinical trials [28] (Supplementary material appendix 2) and TIDieR checklist for the description of the intervention (Supplementary material appendix 3). The study process is outlined in Figure 1.

### Study participants

As this is a feasibility study, we aim to recruit up to 60 service users (PLwD) (with varying levels of Dementia from early diagnosis to moderate stages of Dementia) (30 service users in each group) to test the feasibility, usability and acceptability of KOKU-LITE, feasibility of the recruitment processes, and suitability of the outcome measures. Up to 15 carers, and health and social care professionals (HSCPs) (including General Practitioners (GPs), geriatricians, clinical psychologists, psychiatrists, community psychiatric nurses, physiotherapists, occupational therapists, community nurses, occupational therapists, and music therapists) (or until saturation) will be recruited to evaluate the usability and acceptability of the app for this target population.

### Participant inclusion/exclusion

#### criteria Inclusion criteria

##### PLwD and carers of PLwD

- PLwD aged ≥55 yrs
- willing and able to give informed consent (assessed by the trained researchers and/or HSCPs).
- who lack capacity to consent themselves but can be supported by a carer or an advocate.
- Carers of PLwD who lack capacity to consent themselves.
- able to speak English.
- able to see the tablet/iPad-based app and read instructions with or without glasses.
- able to use tablet/iPad safely, as assessed by the trained research staff and/or HSCPs.
- able to move indoors without help, and with or without a walking aid.

##### Professionals

- HSCPs caring for PLwD in the community

##### Exclusion Criteria

###### PLwD

- Participants with:

- Acute/chronic or uncontrolled medical condition (e.g. severe congestive cardiac failure, uncontrolled hypertension, acute systemic illness, neurological problems, poorly controlled diabetes).
- recent fracture or surgery (within 6 months)
- orthopaedic surgery (such as hip/knee surgery) in the past six months or on a waiting list to have the surgery.
- heart problems such as myocardial infarction or stroke in the past six months
- conditions requiring a specialist/exercise programme (e.g., uncontrolled epilepsy, or uses a wheelchair to mobilise indoors).
- severe hearing/visual impairment that cannot be corrected with aids.
- any other medical condition likely to compromise the ability to use/access the app.
- Participants currently in hospital or care home,
- Participants who have limited understanding or ability to speak English

#### Intervention design

##### Comparator- A Dementia specific falls prevention leaflet

The control group will receive a Dementia specific falls prevention leaflet.

#### Study Intervention – KOKU-LITE program

- Participants will attend a KOKU-LITE group training session lasting for 60 minutes. A carer may also accompany if the participant feels that their support would be beneficial. An iPad will be provided to participants for the duration of the project and the app instruction booklet will also be provided for reference purposes. For participants who do not wish to attend the group training, ad hoc one-to-one training will be offered.
- Participants will be advised to use the modified KOKU-LITE program for 30 minutes, 3 times per week, for 6 weeks.
- During the intervention, the researcher will carry out a weekly visit to participants (either at participants home or a dementia Cafe) based on participants preferences to provide support (if required). In between the sessions, participants will be advised to contact the research team or dementia support advisors (who organise the cafes’) if they experience any difficulties using the app. Contact details of the research team will be provided to the participants.

### Outcome assessment

#### Primary Outcomes

The feasibility and acceptability of the intervention will be evaluated in terms of recruitment, retention and adherence rates to the trial.

#### Secondary Outcomes

Quality of life (QoL) will be measured using The European Quality of Life 5 Dimensions (EQ- 5D-5L) scale. In addition to the QoL, Participants’ Activities of Daily Living (ADL) physical activity levels, functional ability, lower limb strength, fear of falling, falls risk, mood, and user’s experience of the technology (KOKU-LITE app in this study) will be measured. Past medical history (including fractures), medication co-morbidities and demographic data (age, gender, ethnicity, socioeconomic status etc) will also be recorded. Vision will be assessed using a self-reported question, “At the present time, would you say that your eyesight using both eyes (with glasses or contact lenses if you wear them) is” (response set: Excellent, Good, fair, Poor, Very poor, Registered blind). These will be assessed using a group of standardised instruments (Table 1).

**Table 1:** Outcome measures.

| Outcome | Measure |
| --- | --- |
| Quality of life (QoL) | Measured by The European Quality of Life 5 Dimensions (EQ-5D-5L) scale (29). EQ-5D-5L is a 5-dimension version of the original EQ-5D scale. It has two sections: the EQ-5D descriptive system and the EQ visual analogue scale (EQ VAS). The descriptive system comprises five dimensions: mobility, self-care, usual activities, pain/discomfort and anxiety/depression. Each dimension has 5 levels: no problems, slight problems, moderate problems, severe problems and extreme problems. The EQ VAS records the patient's self-rated health on a vertical visual analogue scale where 0 indicates 'worst health you can imagine' and 100 indicates 'the best health you can imagine'. |
| Activities of Daily Living | The Erlangen Test for Activities of Daily Living test (E-ADL-Test) is a valid and reliable instrument for assessing the ADL capabilities of people with Dementia. It includes 4 domains: Pouring a drink, Cutting a piece of bread, Opening a little cupboard, Washing hands (30). |
| Functional mobility | The Short Physical Performance Battery (SPPB) is a validated and reliable instrument to assess functional mobility in people with cognitive impairment/dementia. The SPPB test includes three different domains: walking, sit-to-stand and balance (31). |
| Lower limb strength | Timed up and go (measurement of lower limb strength) |
| Confidence | The Short Falls Efficacy Scale-International (FES-I). The FES-I is a validated and reliable 7-item tool that measures confidence in performing a range of activities of daily living without falling. This scale has recently been modified to maximise its suitability for a range of different languages and cultures (32). |
| Depression | The 5-item Geriatric Depression Scale screening tool for depression in older adults (validated to be as effective as the 15-item GDS for older adults in a wide range of settings (33). |
| Falls risk | Falls risk will be measured with the use of the Assessment of falls risk (FRAT) tool. This validated measure has 5 items which include; history of any fall in the previous year, four or more prescribed medications, diagnosis of stroke or Parkinson's disease, reported problems with balance and inability to rise from a chair without using arms. These are all strong indicators for risk of falling (34). |
| User Experience | The User Experience Questionnaire (UEQ) is a standardized 26-item questionnaire that is used reliably to assess the quality and user experience of the interactive products (35). |

#### Sample Size

The total sample size for this study is 75 (60 service users for the feasibility study and 15 HSCPs for the interviews).

The intended sample size for the feasibility study will be 30 service users in each group (Lancaster et al., 2004) in which simple randomisation will be used with a 1:1 allocation. An attrition rate of 10-20% is reasonable. A formal sample size calculation is not required as this study focuses on the practicalities of conducting the study. This number is pragmatic to assess the feasibility of the KOKU-LITE program with the outcome measurements. It is anticipated that this sample size will provide sufficient information to inform the future definitive trail presuming that the KOKU-LITE program proves feasible and acceptable for PLwD (36).

Up to 15 carers and HSCPs (including General Practitioners (GPs), geriatricians, clinical psychologists, psychiatrists, community psychiatric nurses, community nurses, physiotherapists, occupational therapists, and music therapists) will be recruited to evaluate the usability and acceptability of the app for this target population.

#### Recruitment

-PLwD

#### i) Intervention study

Up to 60 service users (with varying levels of Dementia from early diagnosis to moderate stages of dementia) will be recruited. We will work with professional organisations across Greater Manchester including Dementia United (DU) (DU is Greater Manchester Integrated Care’s programme for dementia), Join Dementia Research (JDR), African Caribbean Care Group (ACCG), Together Dementia support, Alzheimer’s society, and Age UK to identify PLwD who are willing and able to participate in the study. The study information including leaflets and participant information sheet (PIS) will be disseminated on our behalf by DU (established collaboration from previous work), Greater Manchester Mental Health NHS Foundation Trust (GMMH), Age UK, Dementia UK, and other patient organisations to their members and the populations that they serve across Greater Manchester.

In addition, we will use snowball sampling (i.e., participants will be asked to forward the study leaflet and the PIS to individuals who they know of the same condition) to advertise the study and extend our recruitment to individuals who might not be engaged with third sector organisations. Different formats of the PIS will be prepared (if needed) to accommodate the needs of PLwD. For example, different font sizes, and background colours will be used with PIS. Audible versions of the PIS will be provided when required. Participants interested in the study will be advised to contact the research team directly or via their carer (via email) if they are willing to take part in the study. Participants who indicate an interest will then be contacted by the researcher via face-to-face to discuss the study, check eligibility, screen for capacity and answer any questions that participants may have. Assuming participants wish to continue with the study and meet the eligibility criteria, a face-to-face meeting will be arranged either at participant’s home or a Dementia café based on their preference and a time convenient to them. Participants will be sent consent forms in advance of the researcher meeting the participants to ensure that they will have an opportunity to read the consent form and ask any questions that participants may have. Consent forms will be completed at participants’ home or a Dementia café based on their convenience.

#### ii) Interviews

Up to 15 participants (PLwd) who have expressed an interest to participate in the interview will be contacted by the researcher after the completion of the study via face-to-face or email or telephone based on their preferences to discuss the interview part of the study, and answer any questions that participants may have. All participants will be requested to return the completed Consent Form (via email or to the researcher for a face-to-face interview) if willing to participate within one week of receipt. A follow-up attempt will be made for participants failed to respond within one week.

#### iii) Focus groups

Up to 15 carers, and HSCPs (including General Practitioners (GPs), geriatricians, clinical psychologists, psychiatrists, community psychiatric nurses, community nurses, physiotherapists, occupational therapists, and music therapists) will be recruited through personal and professional networks including Royal College of General Practitioners (RCGP), Clinical Research Networks (CRN), JDR, Alzheimer’s Society, Age UK, and GMMH. HSCPs invited to participate in the study will be provided with a PIS and consent form in advance of their involvement in these events. The study information including PIS and consent form will be disseminated on our behalf by the professional organisations. HSCPs who are interested in the study will be advised to contact the researcher, and the researcher will answer any questions the participants may have. All participants will be requested to return the completed Consent Form if willing to participate within 48 hours. A follow-up attempt will be made for participants failed to respond within 48 hours.

#### Additional recruitment strategy

As an additional strategy to supplement recruitment, the researcher will present information about the study at events organised by patient organisations and local carer groups across Greater Manchester to recruit both PLwD and carers. Participants who indicate an interest to take part in the study will complete consent to contact form. Participants who expressed an interest will be contacted by the researcher via face-to-face or email or telephone as preferred to check eligibility, screen for capacity, discuss the study, and answer any questions that participants may have. Assuming participants wish to continue with the study and meet the eligibility criteria, a face-to-face meeting will be arranged either at participant’s home or a Dementia café based on their preference and a time convenient to them. Participants will have one week to consider taking part in the study. A follow-up attempt will be made for participants failed to respond within one week.

#### Randomisation

After assessing for eligibility, baseline assessments and questionnaires will be undertaken and then participants will be randomly allocated to either receive the intervention (KOKU- LITE program+ Dementia specific falls prevention leaflet) or control (Dementia specific falls prevention leaflet). Simple randomisation will be done using sealed envelope (37) (an online platform to perform randomisation). Randomisation will be performed by a researcher independent of the study. Participants will be informed of the randomisation via telephone or email or post. All participants will have an equal chance of being allocated either to the intervention group (KOKU-LITE program) or control group (dementia specific falls prevention leaflet). If participants are allocated to the intervention group, they will use KOKU-LITE for 30 minutes 3 times a week for 6 weeks plus Dementia specific falls prevention leaflet. If participants are allocated to the control group, they will receive dementia specific falls prevention leaflet (and not use the KOKU-LITE intervention).

#### Blinding

Blinding of participants and researchers will not be possible in this study due to the nature of the intervention and due to the lack of resources.

### Data collection

#### Quantitative data

Demographic data and standardised assessments/questionnaires will be completed at baseline and 6 weeks from both the intervention and control groups. Additional questionnaires about usability and motivation to use the app will also be completed at 6 weeks.

* After 6 weeks, a maximum of 15 participants who has expressed an interest will be approached to take part in a semi structured interview to:

a) talk about how the study was carried out and whether improvements could made to the recruitment strategy, PIS etc. and
b) discuss their views about the usability and acceptability of the app. The interview will last approximately 60 minutes.

*Carers, and Health and social care professionals (including General Practitioners (GPs), geriatricians, clinical psychologists, admiral nurses, occupational therapists etc) will be invited to attend up to 3 focus groups, lasting up to 60 minutes. The aim of the focus group is to gather their views about how usable and acceptable the app might be for PLwD, and any changes they would suggest making it suitable for this population.

#### Qualitative research

Semi-structured interviews and focus groups will be conducted either face-to-face or through online platforms (such as MS Teams or Zoom), according to each interviewee’s preference (and in compliance with any restrictions in place due to COVID-19). Interviews or focus groups will last up to 60 minutes. Interview or focus groups will be recorded (audio) with participant’s permission. If an online platform is used, participants will be asked to turn off the video after the researcher and participant introductions. The topic guides used to conduct the interviews will be informed by the literature and our previous work. The topic guide (for service users and carers) will be piloted with the Project Advisory Group (PAG).

### Analysis

#### Quantitative research

Analysis will be descriptive. Baseline characteristics and outcome data will be summarised, as appropriate, using means (with standard deviations) or median (inter-quartile range). For dichotomous or categorical data, frequency will be reported by number or percentage of responses within each category. Calculations will be conducted using SPSS version 24 [check].

#### Qualitative research

The interviews and focus groups will be transcribed verbatim by the research team and then imported into NVivo V. [check] for analysis. Thematic analysis will be undertaken using following the stages outlined in Braun and Clarke (38). The first few interviews (of each group) will be subject to independent coding to achieve consistency.

#### Data monitoring and quality assurance

This study will be subject to the audit and monitoring regime of the University of Manchester. Governmental monitoring agencies may also access study data for monitoring and auditing purposes. A data monitoring committee (DMC) will not be used.

If there is any adverse events or evidence that would cause the researcher to electively stop the research project prematurely, the researchers will make a decision to stop the trial, however, this decision will be based on serious adverse events (SAE). However, no adverse events were reported as a direct result of being involved in the previous KOKU trials. We will stop the trial if the SAE rate in the intervention group is more than 15% higher than the control group.

#### Ethical considerations

The study will be conducted in full conformance with principles of the “Declaration of Helsinki”, Good Clinical Practice (GCP), and within the laws and regulations of the United Kingdom.

The main ethical considerations associated with this project relate to the involvement of PLwD and some of their carers (if of older age) and as such, they are potentially vulnerable. Therefore, the researcher will work closely with patient organisation representatives to ensure that the processes that we put in place are deemed appropriate, suitable and acceptable, and incur minimal participant burden to this vulnerable population.

At least one week before the study initiation, participants will receive PIS describing the study, their voluntary participation and how their personal data & rights will be protected. Informed consent will be obtained before their participation. A distress protocol will be created and adhered to during the study.

##### Informed consent

Capacity to consent is of utmost importance in individuals with Dementia, therefore, we will endeavour to support people with Dementia (PLwD) in consenting for their participation in research. We will adhere to principles of the mental capacity act (39) and the UK Network of Dementia Voices (DEEP) gold standards for dementia research (40). We will abide by the rules and principles of ethical research and informed consent outlined in the DEEP-Ethics gold Standards document recognising for ethical research and informed consent which have been informed by people living with dementia. Written consent will be obtained from all participants. If a person with dementia is unable to provide a signed consent form, then verbal consent will be obtained. Participant information sheets will be developed in easy-read and accessible formats (large font, lay language, short sentences combined with images) if needed. The information sheets will be reviewed by members of the PAG (who are not participants in the study).

##### Patient and public involvement

We have formed a project advisory group (PAG) from the outset for this study. PAG members have lived experiences, either as a person living with dementia or caring for someone with dementia. PAG members will provide support to the research team throughout the study and with interpretation and dissemination of the findings. Patient and public involvement (PPI) will be critical and form an integral part of the project to ensure the project discussions are informed by patients’ and carers’ voices. We will explore the support needed for the participants in terms of feasibility and accessibility of the meetings and documentation for the project including protocol, flyers, participant information sheets, consent forms etc.

The group will continuously provide feedback on documents and meet regularly (at least once a month) to support and provide direction to the research.

##### Data Handling and Management

The rights of the participants will be protected in terms of privacy, confidentiality, pseudo- anonymity, and minimum required information in accordance with GDPR. Access to the participants’ information will be restricted to members of the research team. The data will be pseudonymised (participants’ names will not be used and any identifying information will be removed and replaced with a unique ID number).

Electronic data including audio consent recordings will be held on University of Manchester’s network drives on a password-protected computer owned by the University of Manchester (UoM) accessed from the research team’s own homes (due to hybrid working). Paper-based data will be stored in a digital locked filing cabinet in research team’s office at the University of Manchester. Audio consent forms will be transcribed by the research team. Assessments, questionnaires, signed consent forms, transcripts of audio consent and permission to contact forms will be digitised and stored on the University of Manchester’s secure database on network drives in a password-protected computer as soon as practicable.

##### Dissemination

The results of the study will be disseminated through traditional routes including peer-reviewed publications and at conferences, through patient organisations newsletters and websites, and through social media. To increase the public understanding of the study, the study outputs will be presented at public events organised by dementia-specific patient and third sector organisations.

Impact at international level will be achieved through UoM Applied Research Collaboration (ARC) and KOKU website, which might include descriptions of the study and app prototype, and future work.

##### Discussion

To our knowledge, this is the first trial that explores the feasibility of a gamified digital health program aimed at preventing falls in PLwD in the community. The KOKU-LITE program has been modified by incorporating feedback from patient and public involvement and engagement representatives to suit their needs (supplementary appendix 1). The exercises in the programme will start slowly and progress with the user there by building confidence and reducing fear of falling. Early testing as part of the iterative development process of KOKU-LITE showed that participants valued having the remote facility to exercise and enjoy the health literacy games without the need for the internet. They also found it be accessible and user-friendly. Participants liked the animated video that demonstrated the exercises for them as it motivated them to perform the exercises. Our study will further explore and examine how the KOKU-LITE program can be delivered effectively in the community dwelling PLwD.

We will recruit participants from across the spectrum (including individuals with different types of dementia and different stages of the disease, ethnicity, socio-economic status) to evaluate benefits of the intervention on quality of life and well-being, and to cater a range of physical abilities of PLwD. This will also provide us an opportunity to gain an understanding of the feasibility of the participants with communication difficulties filling out the questionnaires. We will also investigate the feasibility of different outcome measures used in this study which will add to the body of work on outcome measures for this population- this will also aid in the development of new measures if the existing ones are found to be impractical.

Blinding of participants and researchers will not be possible in this study due to the nature of the intervention; however, randomisation will be performed by a researcher independent of the study.

The barriers and facilitators identified in the recruitment and retention phase of the trial will help us to design a robust definitive trial.

##### Ethical approval

This study received favourable ethical approval from Yorkshire & The Humber - Bradford Leeds Research Ethics Committee, Newcastle upon Tyne (REC reference 23/YH/0262)

##### Clinical Trial Registration

The study was registered with the National Clinical Trials Registry with the Unique identifier: NCT06149702.

## Data Availability

At the end of the project, we will deposit a fully anonymised dataset in an open data repository - Figshare at the University of Manchester Library, as well as by request from the primary and corresponding author on reasonable request.

## Author Affiliations

Jaheeda Gangannagaripalli, Research Fellow, Chief investigator, Healthy Ageing Research Group, Division of Nursing, Midwifery and Social Work, Faculty of Biology, Medicine and Health, School of Health Sciences, University of Manchester, Manchester, UK. Chief investigator.

Professor Emma Vardy, Consultant Geriatrician at Northern Care Alliance NHS Foundation trust and Honorary Clinical Chair Manchester Academic Health Sciences network and deputy ageing theme lead for the Greater Manchester NIHR Applied Research Collaboration. Co-investigator

Dr Emma Stanmore, Reader, Healthy Ageing Research Group, Division of Nursing, Midwifery and Social Work, Faculty of Biology, Medicine and Health, School of Health Sciences, University of Manchester, Manchester, UK. Co-investigator

## Author contributions

Study concept and design: ES, EV

Drafting protocol manuscript: JG, ES

Revising the manuscript for important intellectual content; final approval of manuscript, agreement to be accountable for all aspects of the work: JG, ES, EV

## Availability of data and materials

### Funding

This work is supported by the National Institute of Health Research (NIHR) and Alzheimer’s Society (award number: NIHR200174).

