## Supplementary material 1 for "A digital program to prevent falls and improve well-being in people living with dementia in the community: the KOKU-LITE feasibility Randomised Controlled Trial protocol"

**KOKU-LITE supplementary material**

**Appendix 1: Modifications to KOKU program to make it KOKU-LITE**

| **Changes proposed by participants to KOKU program** | **Changes implemented** | **Changes to be implemented** |
| --- | --- | --- |
| Having a video by people living with Dementia to provide feedback/benefits of the app | We have now created a video with one of project advisory group members |  |
| Daily exercises put pressure on people, so having sentence such as – ‘do it at your own pace and at your own comfort’ would be useful. | We have taken this feedback on board, and participants will be advised of this when KOKU-LITE is introduced to participants. | We have not included this in KOKU-LITE program as some participants suggested as ‘LESS is MORE (bearing in mind the condition)’ |
| Having a two-person brain health game that improves social interaction and well-being would be useful. | The research team has now developed the brain health game and is available in the KOKU-LITE program. |  |
| Reduce the questionnaire responses to a minimum for opening questions on falls. | We have removed wellbeing questionnaire (asked at the start using the app, then midway and end of the program) |  |
| Text should be displayed and highlighted along with the animation in different colours. Animation should highlight the part of the body where it is moving. |  | We could not address this change due to funding constraint, however we will introduce these changes in the next version. |
| You might experience pain/discomfort initially when you start this exercise- point out to the specific area where the pain will be. | We will provide this advice as part of the safety advice to keep text to a minimum | We will modify the text and introduce this change in the next version |
| Make it personalised- based on the demographic information the animation should be able to change the voice and name of the person doing the exercise. |  | We could not make this change due to funding constraints; however, we will introduce these changes in the next version |
| Letters are too small on the pictures to recognise | We have now addressed this feedback |  |
| Can't figure out some pictures especially green things and the letters/some pictures are too small to figure out | We have now addressed this feedback |  |
