## Supplementary material 3 for "A digital program to prevent falls and improve well-being in people living with dementia in the community: the KOKU-LITE feasibility Randomised Controlled Trial protocol"

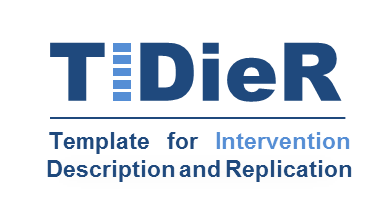
**The TIDieR (Template for Intervention Description and Replication) Checklist*:**

Information to include when describing an intervention and the location of the information

| **Item number** | **Item** | **Where located **** | |
| --- | --- | --- | --- |
|  |  | Primary paper  (page or appendix  number) | Other ^†^ (details) |
|  | **BRIEF NAME** |  |  |
| **1.** | Provide the name or a phrase that describes the intervention. | _____1_______ | A digital program to prevent falls and improve well-being in people living with Dementia in the community: the Keep-On-Keep-Up (KOKU)-LITE feasibility RCT protocol |
|  | **WHY** |  |  |
| **2.** | Describe any rationale, theory, or goal of the elements essential to the intervention. | ______3,6______ | KOKU is a gamified, digital health program designed to maintain function and reduce falls through strength & balance exercises (FaME/OTAGO), and health literacy games including hydration, nutrition, home safety, brain and bone health. KOKU has been modified to suit the needs and preferences of people living with dementia known as KOKU-LITE. This feasibility trial will explore whether KOKU-LITE can be used to support people living with Dementia in the community and to test the trial processes and its implementation. |
|  | **WHAT** |  |  |
| **3.** | Materials: Describe any physical or informational materials used in the intervention, including those provided to participants or used in intervention delivery or in training of intervention providers. Provide information on where the materials can be accessed (e.g. online appendix, URL). | ________4____ | KOKU program can be accessed from the KOKU Health website: <https://kokuhealth.com/>.  **Intervention:** KOKU-LITE program + Dementia-falls leaflet  **Control:** Dementia-falls leaflet  Dementia and falls leaf can be accessed from: <https://www.dementiauk.org/wp-content/uploads/dementia-uk-falls.pdf-> |
| **4.** | Procedures: Describe each of the procedures, activities, and/or processes used in the intervention, including any enabling or support activities. | ___ 4, 6______ | Over a 6-week period, participants will be randomly allocated to either intervention (KOKU-LITE+Dementia specific-falls prevention leaflet) or the control (Dementia-specific-falls prevention leaflet). Participants will be advised to use the KOKU-LITE program for 30 minutes, 3 times per week, for 6 weeks along with the Dementia-falls leaflet (for 6 weeks) and the control group participants will use Dementia falls leaflet for 6 weeks. |
|  | **WHO PROVIDED** |  |  |
| **5.** | For each category of intervention provider (e.g. psychologist, nursing assistant), describe their expertise, background and any specific training given. | ____4,6,9___ | All balance/mobility assessments will be completed by the trained and experienced researchers in the research team. An hour KOKU-LITE training will be provided to participants by the trained and experienced researchers before the data collection commences. |
|  | **HOW** |  |  |
| **6.** | Describe the modes of delivery (e.g. face-to-face or by some other mechanism, such as internet or telephone) of the intervention and whether it was provided individually or in a group. | ____6,________ | Participants in the KOKU-LITE group will attend a KOKU-LITE training session lasting for an hour. An iPad (with KOKU-LITE program) will be provided to participants for 6-weeks for their use at home, and they will be advised to use KOKU-LITE program for 30 minutes, 3 times per week, for 6 weeks. Participants in the control group will be advised to use Dementia-specific-falls prevention leaflet for 6 weeks. |
|  | **WHERE** |  |  |
| **7.** | Describe the type(s) of location(s) where the intervention occurred, including any necessary infrastructure or relevant features. | _____________ | Intervention will be delivered in participants’ homes across Greater Manchester, UK. Participants will be provided necessary training on how to use the KOKU-LITE program, and are advised to use KOKU-LITE program at home. |
|  | **WHEN and HOW MUCH** |  |  |
| **8.** | Describe the number of times the intervention was delivered and over what period of time including the number of sessions, their schedule, and their duration, intensity or dose. | ______6_____ | Participants will be advised to use the KOKU-LITE for 30 minutes, 3 times per week, for 6 weeks along with the Dementia-falls leaflet. During the intervention period, the researcher will visit the participants at least once a week to provide support (if required). |
|  | **TAILORING** |  |  |
| **9.** | If the intervention was planned to be personalised, titrated or adapted, then describe what, why, when, and how. | ______6_______ | The number of home visits to each participant may differ based on their needs and preferences. The exercise program in KOKU-LITE will be tailored to each participant based on their abilities. |
|  | **MODIFICATIONS** |  |  |
| **10.^ǂ^** | If the intervention was modified during the course of the study, describe the changes (what, why, when, and how). | _______N/A______ |  |
|  | **HOW WELL** |  |  |
| **11.** | Planned: If intervention adherence or fidelity was assessed, describe how and by whom, and if any strategies were used to maintain or improve fidelity, describe them. | _______6_____ | Participants in the intervention arm will undergo training on how to use the KOKU-LITE program and a weekly support is available to them based on their preferences. The app instruction booklet will also be provided for reference purposes. Advisors at Dementia Cafes’ have undergone training on how to use the KOKU-LITE program (as part of the KOKU-LITE development) and are available to provide support to the participants (if required).  Adherence: KOKU-LITE program will be tracked using the app; Notifications will be sent to participants to remind them about their next exercise session; and data will be collected at baseline and 6 weeks post-follow-up through validated measures. |
| **12.^ǂ^** | Actual: If intervention adherence or fidelity was assessed, describe the extent to which the intervention was delivered as planned. | ______N/A_______ |  |

** **Authors** - use N/A if an item is not applicable for the intervention being described. **Reviewers** – use ‘?’ if information about the element is not reported/not sufficiently reported.

† If the information is not provided in the primary paper, give details of where this information is available. This may include locations such as a published protocol or other published papers (provide citation details) or a website (provide the URL).

ǂ If completing the TIDieR checklist for a protocol, these items are not relevant to the protocol and cannot be described until the study is complete.

* We strongly recommend using this checklist in conjunction with the TIDieR guide (see *BMJ* 2014;348:g1687) which contains an explanation and elaboration for each item.

* The focus of TIDieR is on reporting details of the intervention elements (and where relevant, comparison elements) of a study. Other elements and methodological features of studies are covered by other reporting statements and checklists and have not been duplicated as part of the TIDieR checklist. When a **randomised trial** is being reported, the TIDieR checklist should be used in conjunction with the CONSORT statement (see [www.consort-statement.org](http://www.consort-statement.org)) as an extension of **Item 5 of the CONSORT 2010 Statement.** When a **clinical trial** **protocol** is being reported, the TIDieR checklist should be used in conjunction with the SPIRIT statement as an extension of **Item 11 of the SPIRIT 2013 Statement** (see [www.spirit-statement.org](http://www.spirit-statement.org)). For alternate study designs, TIDieR can be used in conjunction with the appropriate checklist for that study design (see [www.equator-network.org](http://www.equator-network.org)).
